# Design and context of use of postpartum haemorrhage kits in the UK: a qualitative study combining human factors and social science analysis

**DOI:** 10.1101/2025.09.15.25335093

**Authors:** Matthew Woodward, Alison Powell, Mary Dixon-Woods, Jenni Burt, Cathy Winter, Katherine Lattey, Tim J. Draycott, PPH Kits Contributor Group, Jan W. van der Scheer

**Author notes:** **Correspondence to:** Dr Matthew Woodward, THIS Institute (The Healthcare Improvement Studies Institute), University of Cambridge, Strangeways Research Laboratory, Cambridge CB1 8RN, UK.

## Abstract

Effective management of postpartum haemorrhage (PPH) – a leading cause of maternal mortality – depends on rapid access to critical equipment and medications. PPH emergency kits support timely care, but their design and practical use remain largely unexamined. We combined human factors and social science analysis – drawing on photographs, field observations, and 19 interviews with maternity professionals – to examine kit designs, usability, and contextual factors in six UK maternity units. Many design features, including kit format, item grouping, item visibility, and portability, varied substantially across units. Usability was shaped by factors including labour room and storage constraints, medication access, staff skill mix, and restocking procedures. Some designs risked delays or confusion in emergencies. While standardisation may improve kit safety and efficiency, rigid solutions may risk misalignment with local workflows. A modular approach combining standardised core components with adaptable elements, co-designed with end-users and grounded in ergonomic principles may provide an effective solution.

**Highlights:**

- Emergency kits support timely management of life-threatening postpartum haemorrhage.
- We used human factors and social science to examine kit design and context of use.
- Kit format, configuration, and portability varied across six maternity units.
- Kit design was highly variable, with implications for usability and risk.
- Modular kits may improve safety by combining standardised and adaptable features.

## Introduction

Major primary postpartum haemorrhage (PPH) – defined as blood loss of 1000 mL or more within the first 24 hours after birth – is the leading cause of maternal mortality worldwide, and its incidence is increasing in high-income countries despite advances in obstetric care [1–4]. Causes of PPH are often classified under the “four Ts”: tone (poor uterine contraction), trauma (injury to the genital tract, such as a perineal tear), tissue (retained placenta) and thrombin (clotting abnormalities) [5]. The most common cause is uterine atony, accounting for 70–90% of cases [5, 6]. Effective management of PPH requires a multi-professional response – typically within minutes of onset – using key interventions such as intravenous fluid replacement, medications to contract the uterus (uterotonics), medications to reduce bleeding (e.g. tranexamic acid), or physical techniques such as uterine massage [7–12]. Yet even in high-income countries, the equipment and medications needed to manage a PPH are not always reliably accessible at the time of emergency [13].

Maternity emergency kits have been proposed as a way to align evidence-based practice with reliable medication supply chains at the unit level [14, 15]. Most maternity units in the United Kingdom (UK) rely on such kits to support timely and coordinated care during PPH: pre-prepared collections of medications and equipment for rapid access during an emergency, often accompanied by clinical management algorithms. These kits have typically been developed locally with little formal guidance, contributing to variation in their format, configuration, portability, and ease of restocking – each of which can affect performance in high-pressure situations [15]. Yet their design, usability, and integration into clinical workflows remain largely unexamined [16].

Existing studies of emergency kits, for example in anaesthesia, resuscitation, and paediatrics, have demonstrated the value of applying ergonomic principles to kit designs to improve response times and safety [17–21]. Best practice strategies include the use of clear layout, labelling, and task-based grouping to reduce search time, minimise cognitive load, and support sequence-based working [17–21]. However, studies to date have typically focused on optimising kit design within specific clinical settings and interventions, without fully addressing how variation in clinical environments, workflows, and staffing may affect usability in practice.

Designing emergency kits requires attention not only to the intended clinical interventions, but also to the *contexts of use* – the tasks, users, physical environments, and organisational settings in which they are deployed [22–25]. This principle, well established in human factors and ergonomics, is especially salient in UK maternity care, where settings range from obstetric-led hospital units to freestanding midwifery units located outside a hospital, and vary in infrastructure, staff skill mix, clinical workflows, and medication access. As a result, efforts to improve kit design must balance the benefits of standardisation – such as promoting shared expectations and supporting staff transitions across teams or sites – with the need to accommodate variation across maternity settings [26–29].

Recent work in maternity care has highlighted the value of combining human factors and social science perspectives to address the complexity of healthcare improvement [30], particularly where solutions for systems-level change must be implemented within variable and adaptive settings [31, 32]. Building on previously described methods [16], our study used this interdisciplinary approach to explore the design and context of use of PPH kits across a sample of UK maternity units. Our objectives were to assess the variation in current kit designs, examine how these designs influence usability, understand the context of use of the kits, and identify implications for future design.

## Methods

We drew on photographs, field observations, and interviews with maternity professionals to examine kit designs, usability, and contextual factors in six purposively selected maternity units within the UK National Health Service (NHS). The study was reviewed and approved by a Health Research Authority Research Ethics Committee (reference: 20/NW/0248). Data collection took place between March and October 2023.

### Study setting and participants

We selected a sample of maternity units that reflected variation across the UK in geographic location (different UK regions), size (annual number of births), and unit types (**Table 1)**. Selection was also guided by the units’ availability to participate in the research. The six maternity units included in the study were based in three NHS trusts. In these units, we purposively invited maternity professionals to take part in an interview (**Table 2**). Our sampling focused on those with direct experience of using PPH kits during emergencies or in supporting their use (e.g. retrieving or restocking), while ensuring representation of a range of roles and seniority levels.

**Table 1.** Description of maternity units types in the UK [33].

| Unit type | Description |
| --- | --- |
| Obstetric unit | Hospital-based unit equipped to handle a wider range of pregnancy and birth situations, including those with potential complications or medical needs. On-site access to midwives, obstetricians, anaesthetics, and neonatal services. |
| Alongside midwifery unit | Midwifery-led unit located within or adjacent to a hospital with an obstetric unit. Intended for women with low-risk pregnancies, with access to obstetric, anaesthetic, and neonatal care typically available via a straightforward on-site transfer. |
| Freestanding midwifery unit | Midwifery-led unit located away from a hospital with an obstetric unit. Transfers to specialist care require emergency transport which can take variable amounts of time. |

**Table 2.** Characteristics of included maternity units and interview participants.

| NHS trust* | Size (births per year) | Description of unit/s in trust | Interviewees (N=19) |
| --- | --- | --- | --- |
| 1 | ~ 6,000 | 1 obstetric unit<br>2 freestanding midwifery units | 5 midwives<br>1 labour ward manager (midwife)<br>1 maternity care assistant (MCA) |
| 2 | ~ 5,000 | 1 obstetric unit | 3 midwives<br>1 labour ward manager (midwife)<br>1 maternity care assistant |
| 3 | ~ 3,000 | 1 obstetric unit with alongside midwifery unit<br>1 freestanding midwifery unit | 5 midwives<br>1 labour ward manager (midwife)<br>1 maternity care assistant |

\* NHS trusts are organisational groupings that typically manage multiple hospital and community services within a local area in the UK.

### Data collection

A human factors engineer (MW) and a social scientist (AP) conducted site visits, including making photographs, taking field observations, and conducting semi-structured interviews. The engineer examined the PPH kits in their storage locations, unpacked them to assess internal configuration, and photographed them systematically (typically 12–15 photographs per kit). The engineer’s field observations focused on design characteristics (e.g. kit format and portability), application of ergonomic principles to the kit’s design (**Table 3**), contextual factors of the physical environment affecting kit use (e.g. access to medication, storage constraints, labour room layout), and procedural factors (e.g. restocking and procurement processes). The social scientist focused observations on social and organisational aspects of the context, such as staffing models, task distribution, and training. Observations were supplemented by informal conversations with on-site staff. Field notes were recorded during visits and expanded afterwards.

**Table 3.** Principles used to assess usability of the kits, derived from established human factors and ergonomics principles [24, 34–36].

| Principle | Description of how the principle guided analysis |
| --- | --- |
| Discriminability | The extent to which item grouping and layout supported differentiation between item types and provided visual cues about kit completeness. |
| Visibility | Ease of locating and identifying items, including through clear labelling or transparent containers. |
| Minimalism | Degree to which kits included only essential items, focusing on the minimal range and quantity of items needed for effective task performance. |
| Portability | Ease of transporting and positioning the kit within the clinical environment. Assessed because a single kit was used across multiple locations. |

Interviews were conducted in person or by telephone by one of the two researchers using a semi-structured topic guide. While there was some overlap in interview content, the human factors engineer primarily explored topics related to kit configuration, usability, restocking and procurement processes. The social scientist focused on staff roles, skills and training, and broader organisational and cultural factors affecting kit use. Informed consent was obtained for all interviews. They were audio-recorded, transcribed verbatim by a transcription company, and anonymised prior to analysis.

### Data analysis

Data analysis focused on: (1) mapping the design and configuration of the kits and assessing how specific features shaped their usability, and (2) examining the context of use that influenced how kits were designed, deployed, and maintained.

Photographs and field notes were used to map the design and configuration of each kit, including cataloguing the location of items (e.g. medications, equipment) by container type (e.g. drawer, bag, box). Guided by a high-level task description of PPH management [7, 37], the human factors engineer (MW) conducted a structured usability assessment based on four key principles derived from established human factors and ergonomics principles (**Table 3**). Kits were reviewed and compared against these principles using the photographs, item catalogues, and field notes. Interview transcripts were coded for quotations describing experiences related to these features.

To analyse the context in which PPH kits were used, we drew primarily on interview transcripts, supported by the field notes. Using pattern coding techniques to group similar transcript excerpts into higher-order categories and themes [38], we examined how kit design and usability were shaped by a range of contextual factors. The analysis was informed by an interdisciplinary lens, combining human factors and social science perspectives to explore how kit design and usability interacted with broader systemic conditions (such as staffing models, communication practices, and training culture). Field notes were used to triangulate interview findings and to capture contextual aspects not always verbalised by participants, such as labour room configurations or storage workarounds. NVivo 14 software (QSR International) and ATLAS.ti software were used to code and organise the data.

## Results

Below, we present: (1) an overview of PPH kit designs observed in the six maternity units, (2) an assessment of their usability against ergonomic principles, (3) an analysis of how context of use interacted with design and usability, and (4) design implications based on our analyses.

### Mapping PPH kit designs

The physical formats and internal configurations of the PPH kits varied considerably across the six units (**Table 4**). Kits demonstrated three broad formats: 1) a plastic box, 2) a PPH emergency drawer within a maternity emergency trolley, sometimes supplemented with a separate bag for fluids, or 3) a dedicated multi-drawer trolley for PPH management (see **Figure 1**).

**Figure 1.**
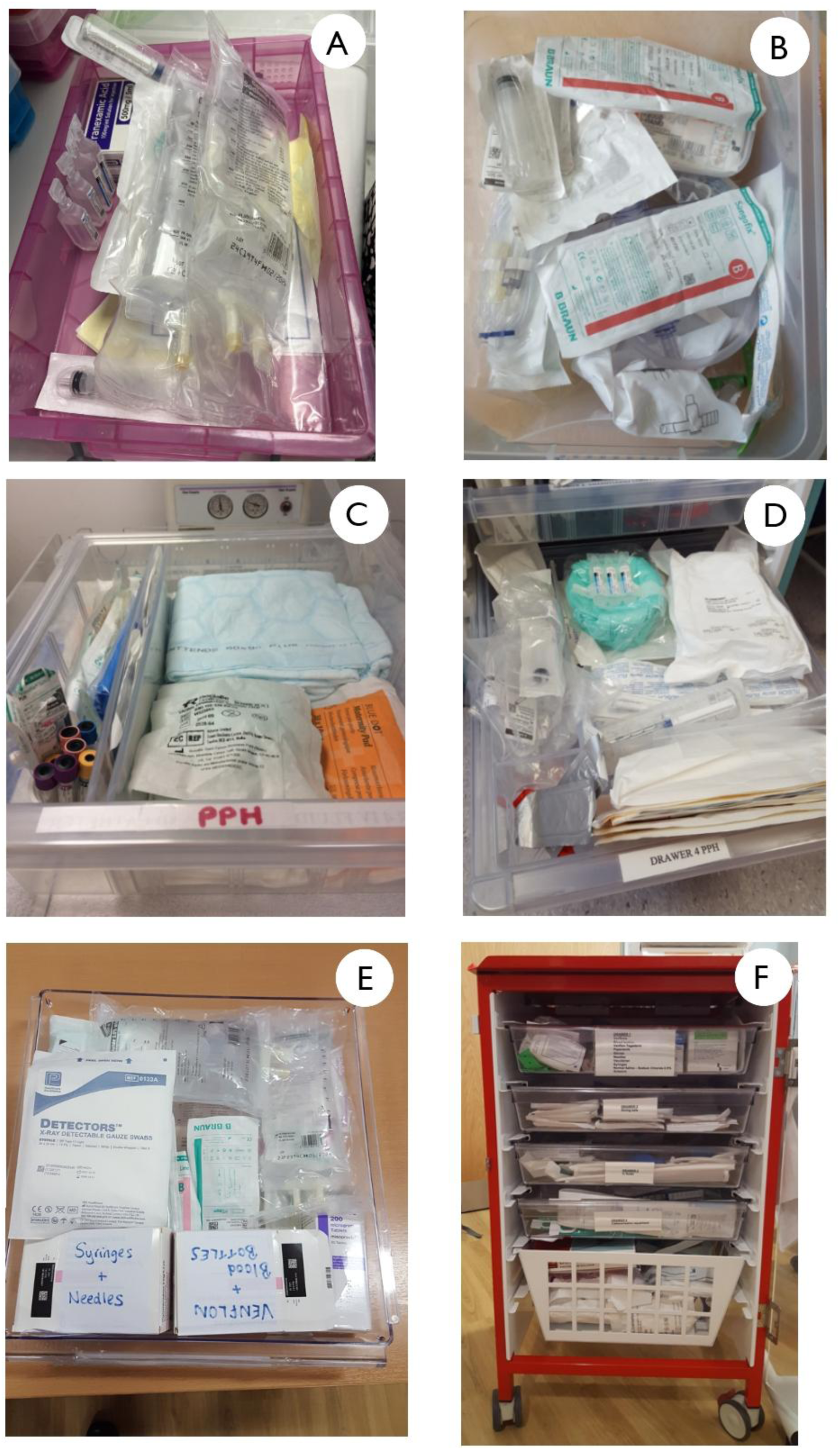
Photographs of the PPH kits used in the six maternity units.

**Table 4.**
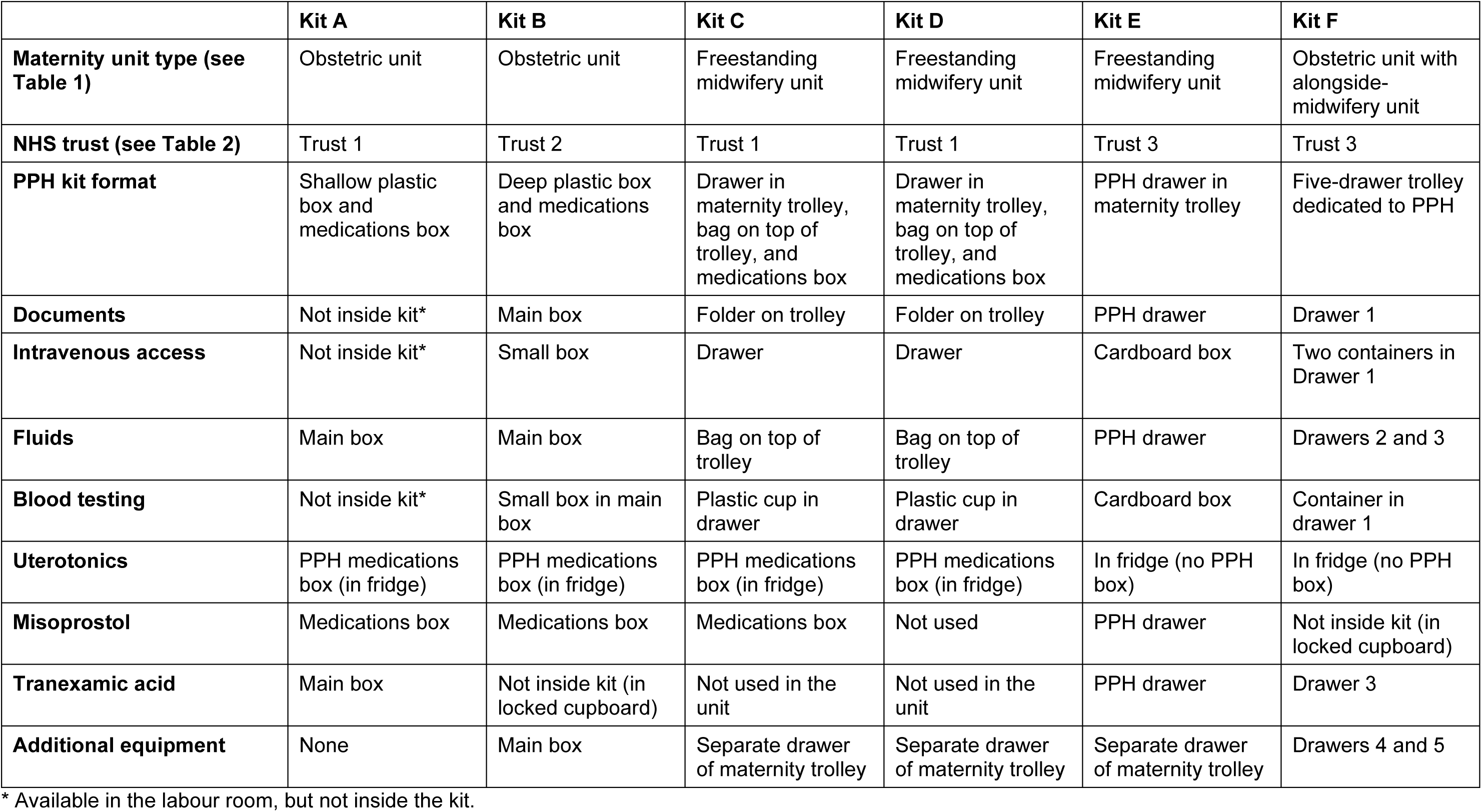
PPH kit designs and configurations across the six units, including a catalogue of the location of items.

|  | **Kit A** | **Kit B** | **Kit C** | **Kit D** | **Kit E** | **Kit F** |
| --- | --- | --- | --- | --- | --- | --- |
| **Maternity unit type (see Table 1)** | Obstetric unit | Obstetric unit | Freestanding midwifery unit | Freestanding midwifery unit | Freestanding midwifery unit | Obstetric unit with alongside-midwifery unit |
| **NHS trust (see Table 2)** | Trust 1 | Trust 2 | Trust 1 | Trust 1 | Trust 3 | Trust 3 |
| **PPH kit format** | Shallow plastic box and medications box | Deep plastic box and medications box | Drawer in maternity trolley, bag on top of trolley, and medications box | Drawer in maternity trolley, bag on top of trolley, and medications box | PPH drawer in maternity trolley | Five-drawer trolley dedicated to PPH |
| **Documents** | Not inside kit* | Main box | Folder on trolley | Folder on trolley | PPH drawer | Drawer 1 |
| **Intravenous access** | Not inside kit* | Small box | Drawer | Drawer | Cardboard box | Two containers in Drawer 1 |
| **Fluids** | Main box | Main box | Bag on top of trolley | Bag on top of trolley | PPH drawer | Drawers 2 and 3 |
| **Blood testing** | Not inside kit* | Small box in main box | Plastic cup in drawer | Plastic cup in drawer | Cardboard box | Container in drawer 1 |
| **Uterotonics** | PPH medications box (in fridge) | PPH medications box (in fridge) | PPH medications box (in fridge) | PPH medications box (in fridge) | In fridge (no PPH box) | In fridge (no PPH box) |
| **Misoprostol** | Medications box | Medications box | Medications box | Not used | PPH drawer | Not inside kit (in locked cupboard) |
| **Tranexamic acid** | Main box | Not inside kit (in locked cupboard) | Not used in the unit | Not used in the unit | PPH drawer | Drawer 3 |
| **Additional equipment** | None | Main box | Separate drawer of maternity trolley | Separate drawer of maternity trolley | Separate drawer of maternity trolley | Drawers 4 and 5 |

\* Available in the labour room, but not inside the kit.

All kits were organised into at least two components: one for items to fulfil tasks such as administering medications and taking blood samples, and one for medications that needed separate storage due to temperature or access controls. **Table 4** outlines how key contents were grouped and stored in various boxes and open containers across the six units, using the following categories:

- Documents – management algorithms and forms to support documentation of clinical actions or restocking
- Intravenous access – cannulas, syringes, tourniquets, and other items needed to establish intravenous access
- Fluids – intravenous fluids and giving sets for volume replacement
- Blood testing – equipment used for taking emergency blood samples
- Injectable uterotonics – vials of medications including oxytocin, ergometrine, Syntometrine®, and carboprost
- Misoprostol – tablet uterotonic
- Tranexamic acid – a medication that works on the clotting pathways to reduce blood loss
- Additional equipment – including urinary catheters, gloves and uterine balloons

### Assessing usability of PPH kits

#### Discriminability

Grouping items into meaningful clusters supports the ergonomics principle of discriminability by helping users distinguish between item categories and reducing search effort [35]. This principle has been applied in the redesign of various emergency kits used in other settings, including anaesthetic and resuscitation trolleys [17, 19, 20, 36, 39], but we found that discriminability (the extent to which the structure of contents supported search) varied significantly across the six PPH kits.

Less structured configurations, such as found in Kits A and B, offered limited support for discriminability. Items were loosely packed in a plastic box with little or no internal organisation. For example, Kit B contained a large number of mixed items (**Figure 1**), with only a small, tightly packed container for intravenous access. Participants described these unstructured configurations as unsatisfactory and the layout was dependent on who last repacked the kit, which had the potential to create uncertainty during emergencies (Kit B in **Figure 1**).

> “I don’t think the box is ideal, and the little box that all the cannula stuff is crammed into. That feels messy when you’re taking it out, I can imagine you’d be scrabbling [Midwife 2, Trust 2]

In contrast, kits with subdivided drawers (e.g. Kits C, D, and F) provided a better internal structure and task-based grouping. For instance, Kit F, a five-drawer dedicated trolley, used drawer separation to group equipment by task from top to bottom, along with grouping within the drawer (**Figure 1**). This organisation reduced the need for searching and enabled parallel working, where different team members could retrieve or use items simultaneously without interference [20]. A similar approach was applied in Kits C and D, where intravenous access and fluid bags were packed in a removable bag.

Across all six units, temperature-controlled medications (e.g. oxytocin, carboprost, and ergometrine) were stored in a fridge (in accordance with manufacturer guidance) and separately from the main kit. These were grouped into a small medication box in four units – in the other two units, the medications were stored loose in the fridge, which risked delay during an emergency, particularly for team members less familiar with the specific kit.

> “The drugs really, I find that quite awful when I’m in and there’s just stuff everywhere…but, to me, I know where stuff is now, but for maybe somebody who doesn’t, they’d be, like, I don’t know where stuff is.” [Midwife 2, Trust 3]

#### Visibility

Several emergency kit redesign studies highlight the benefits of visibility-enhancing features such as clear layout, colour coding, transparent compartments, and labelling [20, 21]. Again, however, we found that visibility varied widely across the PPH kits we assessed and was often suboptimal.

Kits A and B, both using a plastic box format, offered the lowest visibility. Items were piled on top of each other, creating visual clutter and making it difficult to quickly locate or identify supplies (Error! Reference source not found.**1**).

> “The problem sometimes with the box is that it’s not like you open a toolbox and you can see it all laid out there in front of you, you know, it’s layers deep, so you do have to…you know…” [Midwife 1, Trust 2]

Kits C and D used opaque, soft-sided bags to hold fluids and infusion sets and this lack of transparency made it impossible to identify contents quickly. Additional or different visibility issues were found in kits that stored supplies in a single drawer (Kits C, D and F). Items were tightly packed, and limited space constrained layout, often obscuring labels and/or preventing clear identification of specific item types. In all kits, there was little or no use of visual cues like consistent labelling, colour-coded zones, or graphics to direct rapid identification.

Kit F offered the clearest visibility (**Figure 2**). Its wide five-drawer trolley format allowed items to be laid out in a single layer, reducing the need to shift or unpack supplies during use. Each drawer was also labelled, aiding navigation and orientation.

> “What works on the drawers, it is, kind of, like, well the first things that you tend to use are on the top.” [Midwife 2, Trust 3].

**Figure 2.**
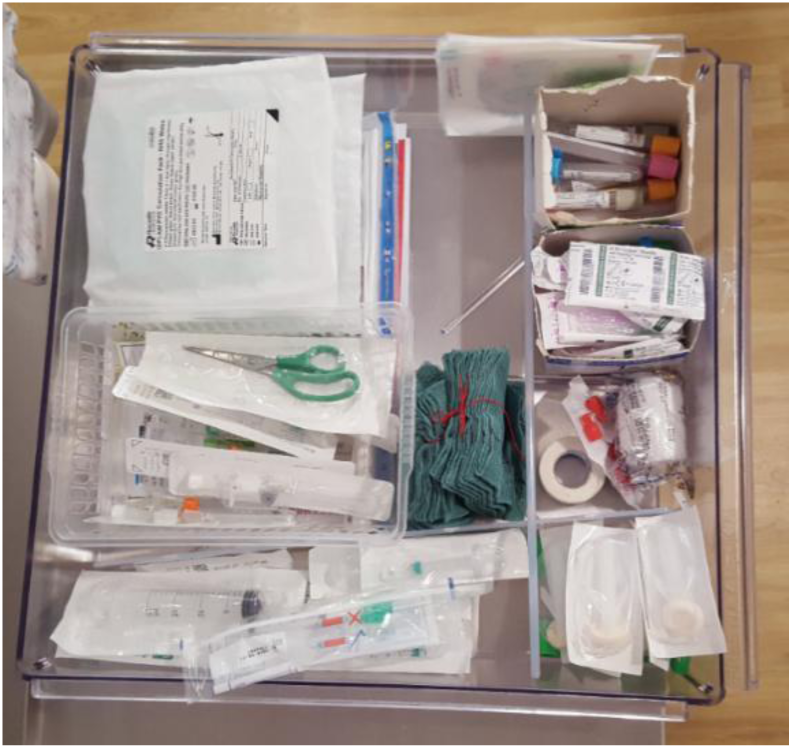
Example of task-based grouping that supports discriminability and visibility (Kit F, drawer 1)

#### Minimalism

Minimalist, streamlined emergency kits enhance usability by reducing cognitive load, improving visibility, and supporting faster item retrieval [21, 34]. We found that the degree to which the PPH kits we examined adhered to the principle of minimalism varied considerably across the six units. Those that were more minimalist tended to be easier to navigate and restock, with fewer items to search through or check for expiry.

Some kits, such as Kits A and C, were intentionally streamlined to include only what was needed for a single PPH emergency (**Figure 3**). This supported task focus and acted as a visual cue for task completion during high-pressure situations.

> “Just making sure that, like, a kit would just only have your essentials in there, otherwise it just gets a bit overwhelming really. Even just one, and just making sure that it gets replaced every time, do you know? You know then that you’ve done each step.” [Midwife 2, Trust 3]

**Figure 3.**
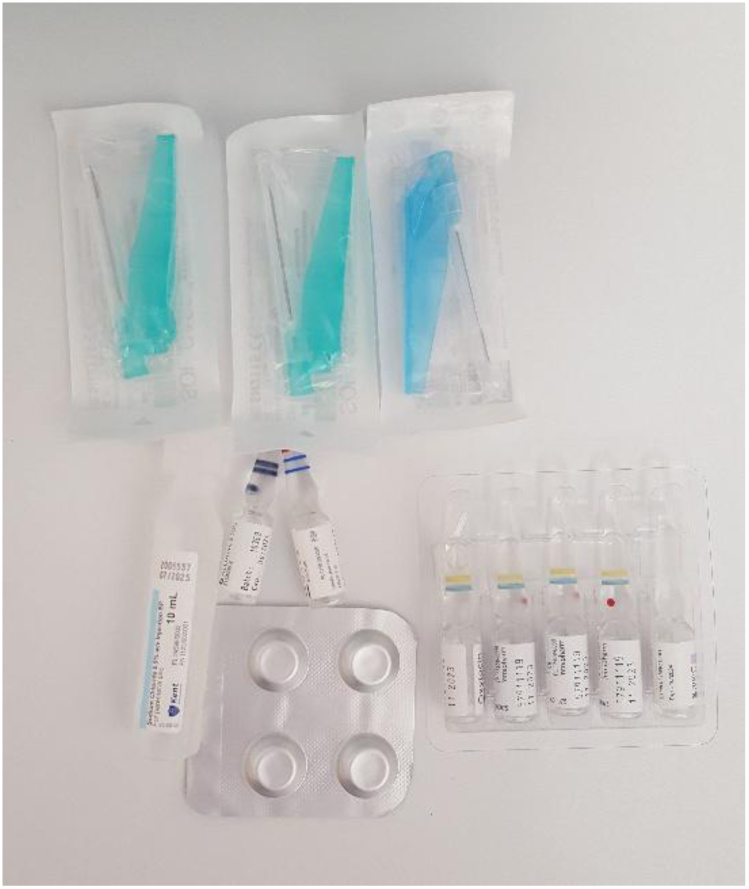
Kit C: Minimalist approach to contents of a medications box (syringes not photographed)

Other kits – most notably the five-drawer trolley (Kit F) – duplicated items. While redundancy offers a buffer if items are missing or faulty, participants described how excessive or non-essential contents (e.g. infrequently used equipment) created clutter, delayed item retrieval, and made kits slower to check.

> “I do feel there’s a lot of, you know, like, so many items that are kept in there and you do… I suppose you could easily miss out one as being out of date.” [Maternity care assistant 3, Trust 3]

While preferences across the midwives and maternity care assistants leaned towards a minimalist approach, in practice most kits included multiples of each item, reflecting a broader tension between the desire for simplicity and the perceived need for preparedness.

#### Portability

The portability of the PPH kits we examined, including their size, shape, and manoeuvrability, varied considerably and had direct implications for workflow efficiency. A single PPH kit was used across multiple locations; therefore, the fit between kit design and the physical environment was an important consideration [23]. Box-format kits (e.g. Kits A and B) were viewed positively for their portability, because they could be carried into the room and placed within arm’s reach of clinicians, minimising movement and reducing time delays. In contrast, trolley-based kits – particularly the five-drawer trolley (Kit F) – were bulky and difficult to manoeuvre easily into tight clinical spaces. Participants noted that during a PPH emergency, labour rooms could become crowded with staff and equipment, limiting the feasibility of bringing in a full trolley. To overcome this challenge, staff sometimes employed workarounds, for example removing a drawer entirely from the trolley to make the kit more portable. These practices underscore how design of the kits interacted closely with their context of use.

### Assessing contexts of use

We identified five key themes that reflect how the design and usability of PPH kits were shaped by their contexts of use: (1) the task of detecting and managing a PPH, (2) the task of checking, restocking, and procuring kit items, (3) users and training, (4) the physical environment, and (5) organisational context. These themes highlight how kit usability was influenced by variations in clinical tasks, staff roles, infrastructure, and local work systems.

#### Detecting and managing a PPH

Although PPH management involves a common core of emergency tasks, participants described how the specific sequence and timing of these tasks depended on assessment of the cause, amount and progression of bleeding, and other clinical characteristics, e.g. body mass index and pre-birth haemoglobin. While guidelines often suggest a linear sequence [7, 37], participants described that tasks were frequently carried out in parallel and dynamically adjusted to the clinical situation.

> “That’s my issue with the PPH proforma [management algorithm] […] it’s so one-size-fits-all, but we all know that there’s different rationales with the bleeding.” [Midwife 4, Trust 1]

Although the most common cause of a PPH is suboptimal uterine tone [5], decisions about administering uterotonics were reported to be influenced by assessments of the current clinical situation and consideration that other causes (e.g. tissue damage) might be responsible for the bleed. Similarly, the threshold for initiating actions like cannulation depended not only on the volume of blood loss but on whether bleeding had been controlled and whether intravenous access was already in place.

#### Checking, restocking and procuring kit items

Kits were checked after each use, with additional routine checks occurring daily or weekly depending on the unit protocol. These checks could be time-consuming – one midwife estimated 20 to 30 minutes – particularly when missing or rarely used items had to be tracked down elsewhere. One unit had implemented a security tag system, which reduced workload by enabling a visual check of a tag code against a logbook to confirm the kit remained intact. Restocking processes and procurement systems varied considerably. Maternity care assistants or midwives usually handled routine checks and replenishment. Basic supplies were typically drawn from centrally managed maternity stockrooms, but ordering processes differed markedly across the units, with some relying on paper-based records and others using electronic medication software. Medications were obtained from the hospital pharmacy, but depending on the unit, either pharmacy technicians or midwives were responsible for stock checks and orders. At one trust, the process and person responsible for external procurement varied depending on the item, for example more specialised devices (e.g. uterine balloons) could only be procured by the ward manager.

#### Users and training

Kits were used by a range of staff with varying roles, responsibilities, and levels of experience and training. Maternity care assistants often fetched the kit during an emergency, retrieved items from it, measured blood loss, and sometimes acted as scribes – meaning they observed and documented the response. Obstetricians often led the clinical response, with anaesthetists usually involved in gaining intravenous access and managing fluid replacement or blood products, but they were less likely to retrieve items directly from the kit. Midwives were typically responsible for drawing up and administering medications, with varying levels of experience depending on whether they typically worked in a high-risk or low-risk setting (where major PPH was less common). Training to use the kits was inconsistent. While some staff received formal instruction, others – especially maternity care assistants – relied heavily on informal, on-the-job learning.

> “[…] it’s very overwhelming, all the information that you have to give because it is not just about what incidents can occur on the ward, but it’s also about what they’re expected to do. […] They do feel very on edge” [Maternity Care Assistant, Trust 2]

Even where structured multi-professional training existed, such as PROMPT emergency training [40], it was variably accessed across and within trusts. Some staff described difficulties securing training slots or felt that centralised sessions were not tailored to the realities of their specific unit.

#### Physical environment

The layout and storage capacity of labour rooms varied significantly across units, with direct impacts on kit contents, placement, and use. In rooms with built-in drawers or cupboards, many supplies were already available at the point of care, allowing for more compact kits. In contrast, rooms with little to no storage required kits to include a broader range of items, increasing their size and complexity. Flat work surfaces such as counters were helpful for setting down kits or drawing up medications, but their availability varied, and in several units, the top of the PPH trolley was occupied with weighing scales or other equipment.

> “We would draw them up, either using one of our delivery trollies, we don’t use the PPH trolley as a rule of thumb for drawing up drugs because […] we use it to measure our blood loss.” [Labour Ward Manager, Trust C]

Due to space constraints and distractions in the labour room, staff often prepared medications elsewhere – typically in the medications/utility room. While supporting cognitive focus, it also introduced time delays, potential communication gaps, and other risks during emergencies.

> “If the room is busy, it can get loud, it can be a little bit…appear chaotic, there is always something going on that everybody knows what they’re doing. It’s often safer to draw them up in the drugs room. However, if you are giving intravenous medication, it should be drawn up in front of the patient that you’re giving it.” [Labour Ward Manager, Trust 3]

Access to temperature-sensitive medications, which were stored in a fridge in an utility room, added time to the PPH response. Thermostable medications, such as misoprostol and tranexamic acid, were variously stored inside kits, in locked cupboards, or separate medications boxes (**Table 4**) – reflecting inconsistency across units.

> “So, on labour ward, it’s a swipe badge access to get into the drugs room but then you need keys to go into the cupboard to get the drugs. That is sometimes problematic finding out which midwife on a large unit has the keys.” [Labour Ward Manager, Trust 1]

These environmental constraints created a trade-off between speed of access and the requirement for secure storage required for certain medications.

#### Organisation

Organisational culture also influenced how PPH kits were configured, maintained, and adapted, consistent with previous evidence on the influence of culture/s on the delivery of care [41, 42]. For example, in some units, a culture of minimalism shaped design choices directly.

> “We’re quite minimalist here, especially our managers, they don’t like clutter, so they like…don’t like it being overstocked.” [Midwife 1, Trust 1]

Variable orientations towards safety and quality improvement were described. In one unit, participants described a receptive environment with strong leadership support for change. In contrast, others described organisational inertia or lack of support. In another unit, one midwife, trying to introduce a patient group direction (PGD) [43] to increase the availability of tranexamic acid (TXA), a medication used in postpartum haemorrhage, reported feeling isolated in the process.

> “Well, I seem to be a bit of a one-man band with it at the moment. So, I think when I went to the last PGD meeting, I think they’ve tried to get TXA as a resource for the [name of unit] before. I don’t know what stopped it.” [Midwife 4, Trust 1]

A practice development midwife at the same trust described how staffing pressures limited opportunities to raise safety concerns or implement improvements. Participants mentioned challenges in modifying established documentation or protocols, such as adapting the management algorithm to reflect diverse settings beyond the obstetric unit. At the same time, participants spoke positively about opportunities for clinical debrief and reflective learning, suggesting that even in resource-constrained environments, there were entry points for fostering improvement.

> “I think our trust is quite good in the range of opportunities that are available to, like, debrief or talk about stuff.” [Midwife 3, Trust 1]

### Design implications

**Table 5** presents implications for improving PPH kit designs, drawing on key insights from the usability assessment and analysis of context of use.

**Table 5.**
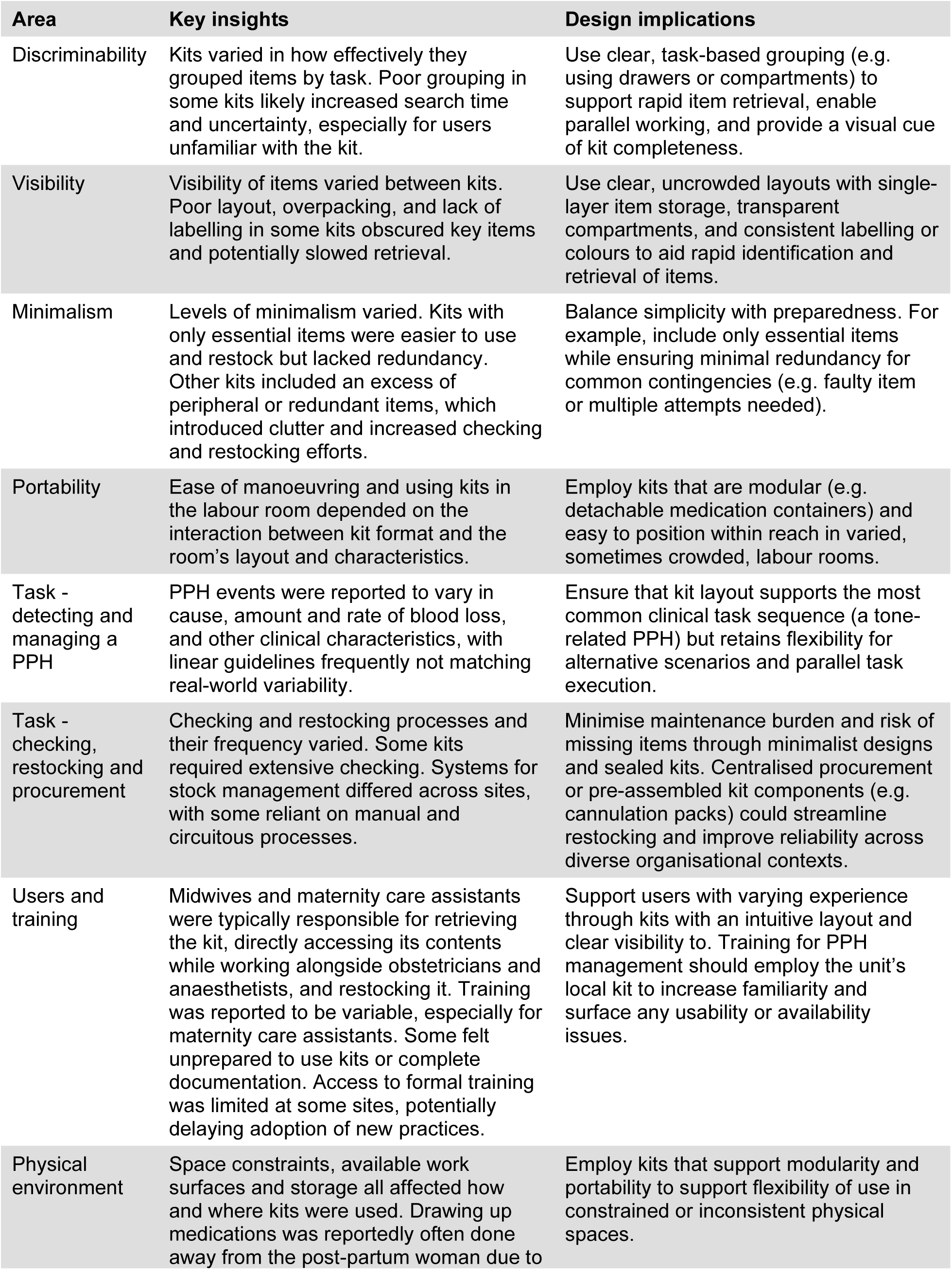
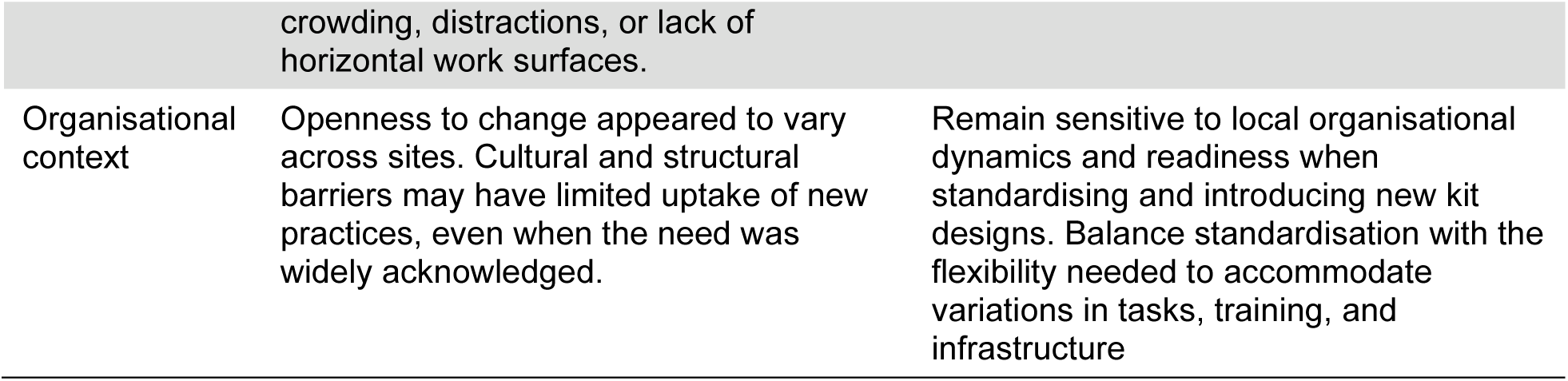
Key insights and design implications drawn from analysis of usability and context of use of PPH kits.

## Discussion

### Main findings

This study identified substantial variation in the design and context of use of PPH kits across UK maternity settings. Kits varied widely in their format and how items were made visible, grouped, and laid out, affecting clinicians’ ability to quickly locate and retrieve critical supplies during an emergency. Important ergonomics principles were applied inconsistently, with several local designs posing risks of delay or confusion in emergency situations. Contextual factors – such as restocking processes, staff training, labour room sizes and setups, access to medications, and improvement capability – also shaped variation in how kits were designed, maintained, and used. We found that uncoordinated local design decisions, in the absence of clear guidance informed by ergonomics principles, may contribute to latent safety risks. These findings highlight the potential for ergonomic improvements to make kits easier and safer to use, and provide evidence of the critical influence of contextual factors on kit design and usability.

### Usability challenges and design implications for current PPH kit designs

This study highlights several usability challenges in the current configuration of PPH kits, underscoring the need for a more systematic application of relevant ergonomics principles – including discriminability, alignment with task sequence, visibility, and minimalism – when designing PPH kits, similar to the redesign of various emergency kits used in other settings, including anaesthetic and resuscitation trolleys [17, 19, 20, 39]. Improvements could include task-based grouping, structuring layout, and selective inclusion of essential items (**Table 5**). Such changes are likely to support timely, reliable clinical responses as well as reducing staff training requirements and simplify kit maintenance.

### Balancing standardisation with local adaptability

Standardisation is widely recognised for promoting shared expectations, enabling rapid action, and facilitating staff transitions across teams or sites [26, 29]. In emergency contexts, standardised kit or equipment layouts have been associated with reduced time to retrieve critical items [18, 44, 45] and may also support procurement and stock management at scale. In time-critical responses, the absence of standardised kit layouts may lead to weakening of shared mental models – especially in maternity care where clinicians regularly move or rotate between different places of work. Similar concerns have emerged in other areas of maternity care, such as early warning systems and fetal surveillance, where a proliferation of non-standard tools has been linked to missed deterioration and delays in escalation [46, 47].

Efforts to standardise PPH kits must navigate a critical tension: while standardisation offers important safety and efficiency benefits, rigid implementation risk misalignment with the varied realities of UK maternity settings. The variation in context of use we identified suggests that a one-size-fits-all approach is unlikely to succeed, as rigid standardisation of PPH kits may risk undermining usability in settings with different infrastructure, storage, or staffing models [48]. When imposed too rigidly, standardisation can create friction, fail to meet local needs, undermine intended outcomes, and struggle to gain traction, [28, 31] unless they accommodate local routines, capacities, and values [31]. This highlights the need for a model of standardisation that is both robust and flexible.

Our analysis supports a modular design approach: combining a standardised core kit – based on the clinical evidence base and containing essential items for initial response tasks such as establishing intravenous access and administering uterotonics – with a flexible, peripheral component that can be tailored to local needs (e.g. whether to include blood weighing scales or catheterisation equipment). This strategy offers a way to retain the benefits of ergonomic design and standardisation to improve emergency preparedness, while accommodating the local realities of varied maternity settings. Future work should mobilise the user-centred design process initiated in this paper, moving from an understanding of context towards co-designing and evaluating prototype solutions with maternity professionals and other relevant users [47, 49, 50].

### Strengths and limitations

Our study highlights how variation in the design and context of use of PPH kits can shape their usability during emergency response. By combining human factors and social science perspectives, we were able to examine both the tangible design features of kits and the physical environment and organisational contexts in which they are deployed. This interdisciplinary approach identified often-overlooked factors that influence how kits are used in practice, including inconsistent restocking processes, ad hoc training for support staff, and the physical constraints of different maternity settings. These findings are consistent with recent calls to integrate diverse methodological lenses in healthcare improvement work, particularly where safety-critical tools are used across variable contexts [25, 30].

Key limitations of this study include a relatively small sample of UK maternity units, which may limit generalisability. For practical reasons, our assessment of usability was based on expert review and interview data rather than usability testing with users. While we did not interview all members of the multi-professional team involved in managing PPH – such as obstetricians and anaesthetists –we focused on midwives and maternity care assistants, who are typically responsible for retrieving the kit, directly accessing its contents, and restocking it. We also did not include home birth settings, since they represent a distinct context with substantially different infrastructure and care models, and were therefore beyond the scope of this study. Despite these limitations, the consistency of themes across diverse unit types, alignment between design observations with user perspectives, and validation by a multidisciplinary authorship team with experience across varied maternity settings lend credibility to the findings and their relevance to wider maternity care contexts.

## Conclusion

The effectiveness and safety of PPH kits in maternity care depends not only on their contents but also on how they are designed, organised, and embedded within local systems. Ergonomic principles and human-centred design provide clear structures to improve usability and mitigate latent safety risks of uncoordinated local design decisions. However, any effort to standardise PPH kits must also be flexible enough to accommodate contextual differences. A promising approach may be to define a modular core kit that supports key emergency tasks, while enabling principled adaptation to local environments and workflows. Our findings underscore the value of integrating human factors and social science insights to understand how work systems shape emergency response.

## Data Availability

The data that support the findings of this evaluation are available upon reasonable request to the authors. &aacute

## Acknowledgements

We’d like to acknowledge the input and guidance of the PPH Kits Contributor Group that included obstetricians, midwives, design specialists and researchers: Arlene Wise, Chloe de Souza, Imogen Brown, Katie Cornthwaite, Lauren Morgan, Louise Swaminathan, Lydia Ufton, Nuala Lucas, Sarah Bell, Sarah Hookes, Sharon Murrell, Sian Harrington, and Steve Summerskill.

## Data availability statement

The data that support the findings of this evaluation are available on request from the corresponding author. Access to fully anonymised data may be granted to bona fide researchers under a data sharing agreement. The data are not publicly available because of privacy or ethical restrictions.

## Contribution to Authorship

Jan W. van der Scheer is the guarantor for this article.

All authors read and approved the final manuscript. Their specific contributions, following <u>CRediT (Contributor Roles Taxonomy)</u>, are as follows:

- Matthew Woodward contributed to conceptualization, data curation, formal analysis, investigation, methodology, project administration, and writing (original draft, review, and editing).
- Alison Powell contributed to formal analysis, investigation, methodology, and writing (original draft, review and editing).
- Mary Dixon-Woods contributed conceptualization, funding acquisition, methodology, supervision, and writing (review and editing).
- Jenni Burt contributed to conceptualization, methodology, and writing (review and editing).
- Cathy Winter contributed to conceptualization, methodology, and writing (review and editing).
- Katherine Lattey contributed to conceptualization, methodology, and writing (review and editing).
- Tim Draycott contributed to conceptualization, methodology, supervision, and writing (review and editing).
- Jan W. van der Scheer contributed to project administration, supervision, writing (original draft, review, and editing).

